# Left Ventricular Remodelling and Functional Status Across Mitral Regurgitation Severity in Mitral Valve Prolapse

**DOI:** 10.64898/2026.09.28.26364239

**Authors:** Klara Lodin, Cristina Da Silva, Anne Wang, Ivana Bulatovic, Lauge Østergaard, Gabriella Boano, Aristomenis Manouras, Lars H Lund, Peter Svenarud, Magnus Dalén, Marcus Carlsson, Jannike Nickander, Bahira Shahim

## Abstract

**Background:** Mitral valve prolapse (MVP) causes mitral regurgitation (MR), leading to volume overload and maladaptive left ventricular (LV) remodelling.

**Objective:** To comprehensively characterize markers of maladaptive LV remodelling across MR severity grades in MVP using cardiovascular magnetic resonance (CMR), and their association with functional status.

**Methods:** Patients with MVP undergoing 1.5T CMR (2016–2025) were stratified by MR fraction: mild<20%, moderate 20–39%, and severe ≥40%. Exclusion criteria were significant coronary artery disease, rheumatic valve disease, mitral stenosis, previous mitral valve procedures, endocarditis or signs of primary cardiomyopathy. Global native T1, T2, extracellular volume (ECV), indexed ECV (iECV), indexed intracellular volume (iICV), and late gadolinium enhancement (LGE) were quantified. Indexed LV end-diastolic volume (EDVI) and the EDV-to-LV mass ratio (EDV/LVM-ratio) were also calculated as indices of LV remodelling and adaptive hypertrophy. Functional status was assessed by NYHA class.

**Results:** Among 112 MVP patients (23 mild, 48 moderate, 41 severe MR) and 20 controls, LGE prevalence (17% vs. 46% vs. 51% vs. 0%) and the proportion with LV dilatation (46% vs. 67% vs. 88% vs 15%) rose progressively with MR severity. Increased EDVI, prevalence of LGE, higher EDV/LVM-ratio, global native T1 and iECV showed the strongest associations with more severe MR, whereas EDV/LVM-ratio, iECV and global native T1 were independently related to NYHA class≥II.

**Conclusions:** CMR markers of maladaptive LV remodelling were abnormal across all MR grades and increased markedly with MR severity. Global native T1, iECV and EDV/LVM-ratio correlated most strongly with symptom burden, rather than MR severity per se.

**Clinical Trial Registration:** NCT06255457

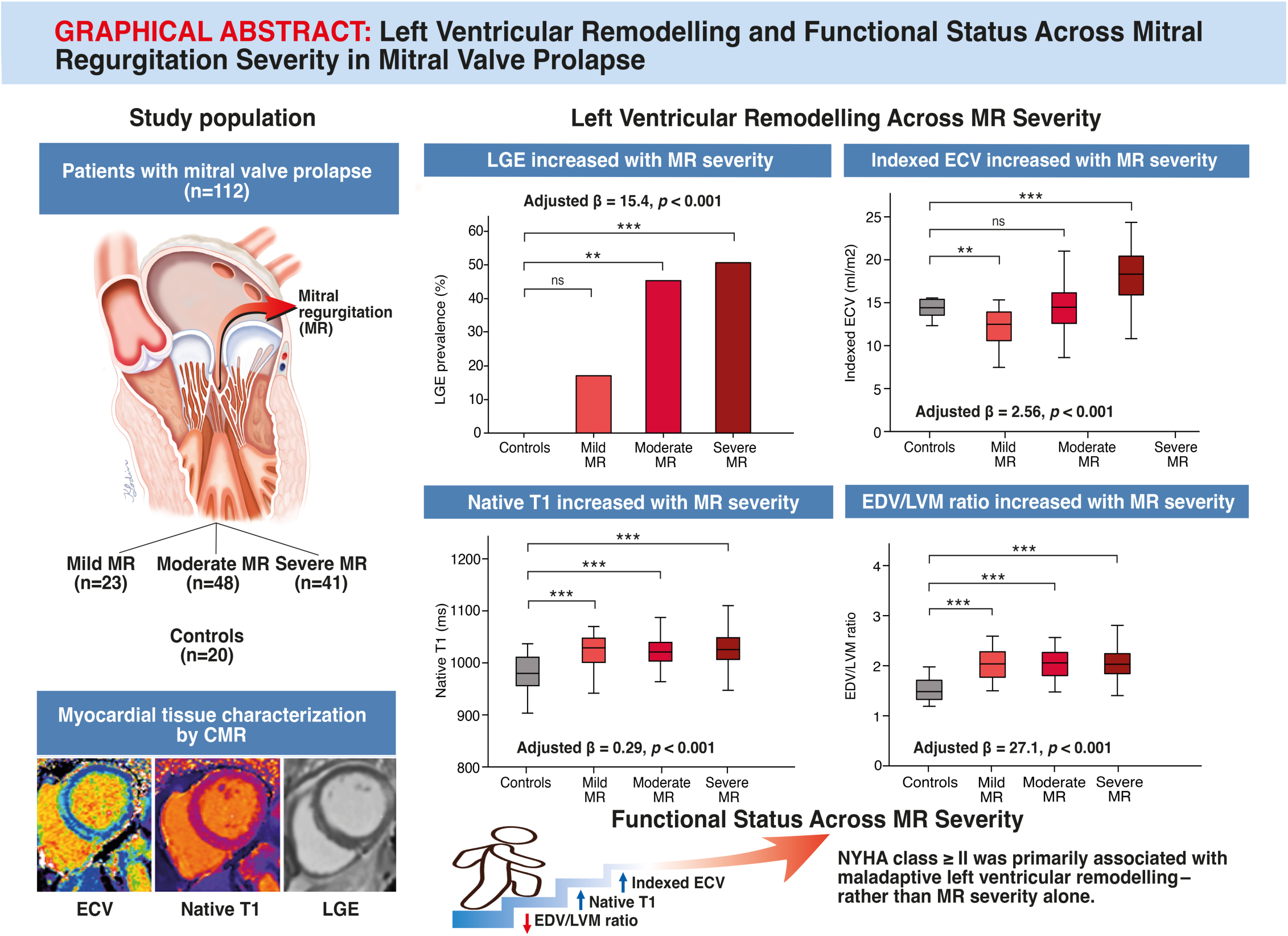

---

Mitral valve prolapse (MVP) is a common valvular abnormality affecting 2–3% of the general population and is the leading cause of primary mitral regurgitation (MR) in developed countries (1). MR in MVP imposes chronic volume overload on the left ventricle (LV), promoting adverse remodelling and, in some patients, heart failure. Cardiovascular magnetic resonance (CMR) imaging has emerged as an alternative modality that provides precise quantification of LV volumes and function, valvular morphology and MR severity, all of which may improve prediction of outcomes and for potential interventions, pre-procedural patient assessment and selection (2, 3). In addition, CMR enables detailed myocardial tissue characterization, with late gadolinium enhancement (LGE) detecting focal, generally replacement, fibrosis, native T1 mapping and extracellular volume (ECV) quantifying diffuse interstitial expansion, generally representing reactive fibrosis, that may precede overt scar (4, 5), and native T2 mapping primarily reflecting myocardial oedema or inflammation (6, 7). Intracellular volume (ICV) further indexes cellular hypertrophy as a component of LV remodelling (8).

Despite this, current criteria for mitral valve surgery in primary MR rely predominantly on echocardiographic grading of MR severity, even though CMR not only provides the above information, but is recognized as more accurate for MR quantification (9). Moreover, by the time patients meet guideline thresholds for “severe” MR, substantial myocardial fibrosis may already have developed, potentially limiting the potential for reverse remodelling and symptomatic improvement (10). Whether earlier intervention guided by markers of myocardial involvement can improve outcomes remains uncertain, emphasising the need to better define structural and tissue changes in earlier stages of MR and to relate these to clinical status.

The present study therefore aimed to provide a comprehensive analysis of quantitative CMR markers of maladaptive LV remodelling across the full spectrum of MR severity in MVP, to compare these findings with controls without MVP or MR, and to evaluate their association with functional status.

## Methods

### Study Population

The study cohort consisted of patients with MVP and mild, moderate or severe degenerative MR and healthy controls without MVP or MR who had performed CMR between June 2016–April 2025 at Karolinska University Hospital, Stockholm, Sweden. Patients with MVP were included from the ongoing MitraVT study (NCT06255457), which enrolls patients with MVP across a range of MR severity, including patients referred for mitral valve surgery due to significant MR as well as patients without an indication for surgery, and investigates associations between CMR-derived biomarkers and clinical outcomes. Non-surgical MVP patients were recruited at the outpatient clinic at Karolinska University Hospital when referred for evaluation of mitral valve disease. Historical controls without MVP were recruited through public advertisements and participated on a voluntary basis and underwent CMR between June 2016 and February 2022 (11, 12). Controls were matched with MVP patients based on age and sex. Among both patients and controls, those with significant coronary artery disease (ruled out by either coronary computed tomography or coronary angiography), rheumatic valve disease, mitral stenosis, previous mitral valve procedures, endocarditis or evidence of primary cardiomyopathy were excluded.

Data on patient demographics and comorbidities were collected from electronic medical records. New York Heart Association (NYHA) class was assessed by the patient’s responsible physician based on patient-reported symptoms. For all patients both echocardiography and CMR examinations were available. Ethical approval was granted by the Swedish Ethical Review Authority.

### Image Acquisition

CMR was performed at 1.5T (MAGNETOM Aera or Sola scanner, Siemens Healthineers, Forchheim, Germany). Full coverage retrospective ECG-gated balanced steady state free precession (SSFP) cine imaging was acquired in short-axis and three long-axis slices.

Three or five short-axis native T1 maps were obtained using an ECG-gated modified look-locker inversion 5s(3s)3s recovery research sequence. Three or five short-axis native T2 maps were acquired, using a T2-prepared sequence (MyoMaps, Siemens Healthineers, Forchheim, Germany). A bolus of total 0.15 mmol/kg gadobutrol (Gadovist or Pixxoscan) was injected intravenously, and LGE images were acquired 10-15 minutes after the injection of contrast media using a free free-breathing phase-sensitive inversion recovery sequence with balanced steady-state free precession single-shot readout. Three or five ECV-maps at rest were generated from native T1-maps and post-contrast T1-maps and calibrated by the hematocrit.

### Image Analysis

Cine images, LGE images, ECV-maps, native T1- and native T2-maps were analysed with the software Segment (version 4.1.0.1 Medviso AB, Lund, Sweden) by carefully delineating the endo- and epicardial borders of the LV in the short-axis images. To further avoid contamination from blood pool and adjacent tissues, a 10% erosion margin was set for both endo- and epicardial borders for the export of the respective mapping values. LV volumes and mass were indexed to body surface area calculated with the Mosteller formula (13). MR volume was calculated as LV stroke volume from cine images delineated in end-diastole and end-systole subtracted by aortic forward flow volume, and the MR fraction was calculated as MR volume divided by LV stroke volume. MR severity was defined by MR fraction, according to previously proposed CMR-specific thresholds for primary MR, and was categorized as mild (<20%), moderate (20–39%), and severe (≥40%) (5).

LV dilatation was defined as increased LV end-diastolic volume index (EDVI) defined as values exceeding the age- and sex-specific upper reference limits derived from CMR reference ranges for smooth segmentation from the Healthy Hearts Consortium (14). The LV end-diastolic volume to LV mass ratio (EDV/LVM-ratio) was calculated and used as a measure of the extent of adaptive LV hypertrophy and thus a reflection of global end-diastolic wall stress (15).

Focal fibrosis was visually assessed in LGE images. Identification of focal fibrosis required confirmation in two contiguous short-axis slices or one short-axis slice and one additional orthogonal slice. To focus on the association of MVP and LV focal fibrosis, and to minimize other confounders, we excluded LGE isolated to right ventricular insertion sites, as this pattern may reflect pulmonary hypertension and right ventricular remodelling that are consequences of severe MR. Global native T1, native T2, and ECV values were acquired by averaging all segments of a 16-segment model of the LV. Indexed extracellular (iECV) and intracellular volume (iICV) were calculated as iECV = ECV × LVM index (LVMI) / 1.05 and iICV = (1 − ECV) × LVMI / 1.05, respectively (8). Global native T1, native T2 and ECV values included foci of nonischaemic scar on LGE images (2, 16).

### Assessment of Mitral Valve Characteristics

MVP was diagnosed according to current guidelines in the parasternal long-axis view by observing a systolic displacement of the mitral leaflet into the left atrium (LA) of at least 2 mm from the mitral annular plane (17). The MVP phenotype was subdivided as single-leaflet (anterior or posterior) or bileaflet prolapse. Subsequently, MVP was further stratified as either Barlow’s disease or “other ethiology”, including fibroelastic deficiency (FED), FED+, and forme fruste. Mitral annular disjunction (MAD) was defined as the distance between the insertion point of the posterior mitral leaflet and the LA–LV junction. Inferolateral MAD was primarily assessed in parasternal long-axis and apical three-chamber views, whereas inferior MAD was best visualized in apical two-chamber or basal short-axis planes. MAD was considered present only if visualized during both systole and diastole, with careful distinction from pseudo-MAD (18). Although CMR is the reference standard, subtle features of MAD and Barlow disease can still be challenging to assess on CMR alone. Therefore, echocardiograms were systematically reviewed alongside CMR studies to reduce the risk of missed abnormalities and to strengthen the diagnostic classification. Transthoracic and transesophageal echocardiograms were performed a median of 79 days (IQR 45−180) days before the CMR examination (Vivid E95 scanner, GE Healthcare and Philips).

### Statistical Analysis

Among patients with MVP, MR severity was treated as an ordinal variable. Categorical variables were presented as percentages and absolute numbers and were compared by Cochran-Armitage test for trend by MR severity. Continuous data were expressed as mean values and standard deviation. Continuous data were compared by Jonckheere-Terpstra test. For comparisons with healthy controls, each MR severity group was analysed separately, with continuous variables compared using independent samples t-tests or Mann–Whitney U tests, as appropriate, and categorical variables analysed using Chi-squared or Fisher’s exact tests. The same approach was applied in the subgroup analysis of patients with MVP referred for mitral valve surgery, comparing asymptomatic patients (NYHA class I) with symptomatic patients (NYHA class ≥ II). Linear regression was performed among patients with MVP to identify characteristics associated with MR severity, with MR fraction entered as the dependent variable. Each imaging parameter was entered separately into the multivariable models adjusted for age, sex, body mass index and hypertension. The results of the linear regression were presented as unstandardized coefficients (β), along with 95% confidence intervals (CI) and p-values. Binary logistic regression analyses were conducted among MVP patients to identify characteristics associated with LV dilatation and NYHA class ≥ II. Each imaging parameter was entered separately into individual multivariable models adjusted for age, sex, body mass index and hypertension. The results of the binary logistic regressions were presented as odds ratios (ORs) with 95% CI and p-values. All tests were two-tailed and p-values < 0.05 were considered significant. Statistical analyses were performed using IBM SPSS Statistics® (IBM SPSS Statistics 28, IBM, New York, USA).

## Results

### Study Population

The study included 20 healthy controls and 112 patients with MVP, stratified by MR severity into mild (n=23), moderate (n=48), and severe (n=41) MR (Table 1). Women were predominantly in the mild MR group and men in the severe MR group. The prevalence of hypertension, diabetes, and hyperlipidemia was overall low and similar between patients and controls. Pulmonary hypertension (resting systolic pulmonary artery pressure >50 mmHg) and atrial fibrillation were more frequent in moderate and severe MR compared with controls (pulmonary hypertension: 0% vs. 15% and 27%; atrial fibrillation: 0% vs. 27% and 22%, respectively). Most patients with mild MR were in NYHA class I, whereas moderate and severe MR included a higher proportion of patients in NYHA class II–III.

**Table 1:** Baseline characteristics stratified by degree of MR in patients with MVP and controls.

| Clinical variables | Controls<br>(n=20) | Mild MR<br>(n=23) | Moderate<br>MR (n=48) | Severe MR<br>(n=41) | p-value<br>Mild vs.<br>moderate<br>vs. severe* | p-value<br>Control<br>vs. mild<br>MR** | p-value<br>Control vs.<br>moderate<br>MR** | p-value<br>Control<br>vs. severe<br>MR** |
| --- | --- | --- | --- | --- | --- | --- | --- | --- |
| Sex, female | 55% (11) | 75% (18) | 52% (25) | 27% (11) | <0.001 | 0.16 | 0.83 | 0.03 |
| Age, years | 52.2±13.2 | 46.0±16.6 | 53.7±17.0 | 55.7±13.7 | 0.06 | 0.20 | 0.62 | 0.23 |
| BMI, kg/m<sup>2</sup> | 24.1±3.5 | 22.7±4.0 | 23.3±3.4 | 23.7±3.4 | 0.22 | 0.20 | 0.49 | 0.69 |
| BSA, m<sup>2</sup> | 1.87±0.20 | 1.79±0.20 | 1.88±0.19 | 1.96±0.22 | 0.002 | 0.17 | 0.90 | 0.13 |
| Hematocrit, % | 42.6±4.0 | 42.8±7.0 | 42.0±3.4 | 42.2±4.3 | 0.99 | 0.74 | 0.60 | 0.77 |
| eGFR,<br>ml/min/1.73m<sup>2</sup> | 79.8±8.9 | 84.5±13.6 | 75.2±16.3 | 78.7±11.0 | 0.18 | 0.25 | 0.50 | 0.74 |
| Creatinine, µmol/l | 74.6±13.4 | 68.4±14.0 | 80.3±16.9 | 77.4±12.6 | 0.04 | 0.14 | 0.39 | 0.61 |
| NTproBNP, pg/ml | — | 150.8±88.5 | 433.5±682.2 | 459.3±724.3 | 0.42 | — | — | — |
| Hypertension | 15% (3) | 13% (3) | 23% (11) | 22% (9) | 0.43 | 1.00 | 0.53 | 0.73 |
| Hyperlipidaemia | 10% (2) | 13% (3) | 2% (1) | 10% (4) | 0.92 | 1.00 | 0.21 | 1.00 |
| Diabetes | 5% (1) | 0% (0) | 2% (1) | 2% (1) | 0.50 | 0.46 | 0.51 | 1.00 |
| Pulmonary<br>hypertension | 0% (0) | 0% (0) | 15% (7) | 27% (11) | 0.004 | — | 0.10 | 0.01 |
| Atrial fibrillation | 0% (0) | 4% (1) | 27% (13) | 22% (9) | 0.16 | 1.000 | 0.01 | 0.02 |
| Stroke | 0% (0) | 0% (0) | 4% (2) | 2% (1) | 0.67 | — | 1.00 | 1.00 |
| Smoking status |  |  |  |  | 0.44 | 0.29 | 0.045 | 0.21 |
| No smoking | 65% (13) | 79% (19) | 88% (42) | 78% (32) |  |  |  |  |
| Previous | 35% (7) | 21% (5) | 13% (6) | 17% (7) |  |  |  |  |
| Current | 0% (0) | 0% (0) | 0% (0) | 5% (2) |  |  |  |  |
| NYHA class |  |  |  |  | 0.001 | — | — | — |
| I | N/A | 79% (19) | 52% (25) | 39% (16) |  |  |  |  |
| II | N/A | 21% (5) | 46% (22) | 54% (22) |  |  |  |  |
| III | N/A | 0% (0) | 2% (1) | 7% (3) |  |  |  |  |
| IV | N/A | 0% (0) | 0% (0) | 0% (0) |  |  |  |  |
| Mitral valve<br>characteristics |  |  |  |  |  |  |  |  |
| MAD inferolateral | N/A | 29% (7) | 27% (13) | 15% (6) | 0.14 | — | — | — |
| MAD inferolateral<br>length, mm | N/A | 7.9±2.0 | 10.0±2.4 | 9.7±3.8 | 0.13 | — | — | — |
| MAD inferior | N/A | 26% (6) | 29% (14) | 17% (7) | 0.32 | — | — | — |
| MAD inferior<br>length, mm | N/A | 6.7±1.8 | 10.0±2.4 | 10.0±3.8 | 0.22 | — | — | — |
| Pseudo-MAD | N/A | 17% (4) | 21% (10) | 10% (4) | 0.35 | — | — | — |
| Barlow's disease | N/A | 54% (13) | 52% (25) | 39% (16) | 0.20 | — | — | — |
| Other etiology | N/A | 46% (11) | 48% (23) | 61% (25) | 0.20 | — | — | — |
| Posterior leaflet<br>prolapse | N/A | 38% (9) | 42% (20) | 66% (27) | 0.02 | — | — | — |
| Anterior leaflet<br>prolapse | N/A | 4% (1) | 8% (4) | 0% (0) | 0.28 | — | — | — |
| Bileaflet prolapse | N/A | 58% (14) | 50% (24) | 34% (14) | 0.048 | — | — | — |
| CMR finding |  |  |  |  |  |  |  |  |
| Heart rate, bpm | 67.8±6.5 | 63.3±10.3 | 65.0±11.9 | 70.5±12.6 | 0.07 | 0.10 | 0.12 | 0.27 |
| LVM, ml | 104.1±23.9 | 84.5±19.5 | 103.1±25.4 | 131.8±37.2 | <0.001 | 0.01 | 0.77 | 0.004 |
| LVM, g | 109.3±25.0 | 88.7±20.5 | 108.2±26.6 | 138.4±39.0 | <0.001 | 0.01 | 0.77 | 0.004 |
| **LVMI, g/m<sup>2</sup>** | 58.0±9.0 | 49.1±8.9 | 57.3±10.0 | 69.7±14.1 | <0.001 | 0.002 | 0.78 | 0.001 |
| **EDV, ml** | 168.0±49.3 | 171.6±37.5 | 206.1±47.0 | 265.8±76.1 | <0.001 | 0.79 | 0.01 | <0.001 |
| **EDVI, ml/m<sup>2</sup>** | 88.7±19.4 | 95.6±18.2 | 109.7±20.2 | 134.7±31.5 | <0.001 | 0.24 | <0.001 | <0.001 |
| **ESV, ml** | 74.3±22.6 | 80.7±22.8 | 90.7±30.5 | 107.7±40.2 | 0.002 | 0.37 | 0.03 | <0.001 |
| **ESVI, ml/m<sup>2</sup>** | 39.3±9.6 | 44.4±10.5 | 48.1±14.1 | 54.4±18.2 | 0.02 | 0.11 | 0.02 | <0.001 |
| **SV, ml** | 93.8±30.0 | 89.6±17.8 | 115.4±26.2 | 159.3±43.0 | <0.001 | 0.57 | 0.004 | <0.001 |
| **SVI, ml/m<sup>2</sup>** | 49.5±12.0 | 50.1±8.8 | 61.6±12.5 | 80.7±17.7 | <0.001 | 0.85 | <0.001 | <0.001 |
| **EF, %** | 55.7±5.4 | 52.8±6.2 | 56.5±8.0 | 60.4±6.6 | <0.001 | 0.11 | 0.70 | 0.01 |
| **Cardiac index, l/min/ m<sup>2</sup>** | 3.6±0.56 | 2.8±0.61 | 2.7±0.55 | 2.7±0.71 | 0.15 | <0.001 | <0.001 | <0.001 |
| **Aorta flow forward, ml** | 88.7±20.1 | 81.3±17.0 | 83.4±19.3 | 78.9±23.0 | 0.39 | 0.23 | 0.37 | 0.15 |
| **LV dilatation** | 15% (3) | 46% (11) | 67% (32) | 88% (36) | <0.001 | 0.03 | <0.001 | <0.001 |
| **EDV/LVM-ratio** | 1.53±0.23 | 2.04±0.32 | 2.03±0.30 | 2.05±0.38 | 0.79 | <0.001 | <0.001 | <0.001 |
| **MR fraction, %** | N/A | 10.6±7.1 | 28.5±6.0 | 50.4±9.1 | <0.001 | — | — | — |
| **MR volume, ml** | N/A | 9.8±7.0 | 33.4±10.5 | 81.5±29.8 | <0.001 | — | — | — |
| **LGE prevalence** | 0% (0)<sup>a</sup> | 17% (4) | 46% (21)<sup>a</sup> | 51% (21) | 0.01 | 0.15 | 0.001 | <0.001 |
| **LGE, ml** | 0.0±0.0 | 0.49±1.1 | 1.4±2.0 | 2.1±2.8 | 0.003 | 0.40 | 0.002 | <0.001 |
| **LGE, %** | 0.0±0.0 | 0.68±1.6 | 1.5±2.1 | 1.8±2.3 | 0.01 | 0.40 | 0.002 | <0.001 |
| **Global Native T1, ms** | 982.6±34.1 | 1022.0±34.7<sup>b</sup> | 1019.1±28.9<sup>b</sup> | 1026.1±32.5<sup>b</sup> | 0.22 | <0.001 | <0.001 | <0.001 |
| **Global Native T2, ms** | 49.1±2.8 | 50.2±3.0<sup>c</sup> | 49.2±1.9<sup>c</sup> | 49.0±2.4<sup>c</sup> | 0.53 | 0.28 | 0.95 | 0.85 |
| **Global ECV, %** | 27.0±2.2 | 26.7±2.7<sup>d</sup> | 27.0±2.6<sup>d</sup> | 27.0±2.7<sup>d</sup> | 0.61 | 0.72 | 0.92 | 0.79 |
| **ECV inferolateral, %** | 25.8±2.9 | 25.7±3.4 | 26.7±4.1 | 28.7±4.8 | 0.01 | 0.87 | 0.48 | 0.03 |
| **iECV, ml/m<sup>2</sup>** | 14.8±2.2 | 12.4±2.6 | 14.8±3.2 | 18.0±3.6 | <0.001 | 0.001 | 0.91 | <0.001 |
| **iICV, ml/m<sup>2</sup>** | 40.4±6.9 | 34.0±6.6 | 39.8±6.9 | 49.0±10.5 | <0.001 | 0.01 | 0.75 | 0.001 |

\*p-value for trend across MR severity groups assessed by Jonckheere-Terpstra test for continuous variables and Cochran-Armitage test for categorical variables.
\*\*p-value for comparison between controls and each MR severity group assessed by independent samples t-test or Mann-Whitney U test for continuous variables and Chi-squared or Fisher's exact test for categorical variables.
Abbreviations: BMI – body mass index, BSA – body surface area, CO – cardiac output, ECV – extracellular volume, EDV – end diastolic volume, EDVI – end diastolic volume index, EF – ejection fraction, ESV – end systolic volume, ESVI – end systolic volume index, iECV – indexed extracellular volume, iICV – indexed intracellular volume, LV – left ventricular, LGE – late gadolinium enhancement, LVM – left ventricular mass, LVMI – left ventricular mass index, MAD – mitral annular disjunction, MR – mitral regurgitation, NYHA – New York Heart Association, SV – stroke volume, SVI – stroke volume index.
<sup>a</sup>LGE images were not acquired in 5 controls and 2 patients with moderate MR.
<sup>b</sup>Native T1 maps were not acquired in 1 patient with mild MR and 1 patient with moderate MR.
<sup>c</sup>Native T2 maps were not acquired in 9 patients with mild MR, 13 patients with moderate MR and 6 patients with severe MR.
<sup>d</sup>ECV maps were not acquired in 4 patients with mild MR, 2 patients with moderate MR and 2 patients with severe MR.

Patients with severe MR more frequently had posterior leaflet prolapse (66%) compared with those with mild (38%) and moderate MR (42%) (p=0.02), whereas anterior leaflet prolapse was rare overall and absent in severe MR. Bileaflet prolapse was more common in mild and moderate MR (58% and 50%, respectively) compared with severe MR (34%) (p=0.048). There were no significant differences in the prevalence of Barlow’s disease or MAD across the MR grade.

### LV Remodelling Across MR Severity

The MR fraction was 10.6±7.1% in mild MR, 28.5±6.0% in moderate MR and 50.4±9.1% in severe MR (p<0.001) (Table 1). Compared with healthy controls, MVP patients showed progressively more pronounced LV remodelling with increasing MR severity. Severe MR was associated with larger LV volumes (EDVI 134.7±31.5 ml/m² vs. 95.6±18.2 ml/m² in mild MR, p<0.001) and higher indexed stroke volumes (80.7±17.7 ml/m² vs. 50.1±8.8 ml/m² in mild MR, p<0.001). LV dilatation was uncommon in controls (15%) but was increasingly prevalent with MR severity (46% in mild MR to 88% in severe MR, p<0.001). LVMI also increased with increasing MR severity (p<0.001). In contrast, cardiac index was lower in patients with mild, moderate, and severe MR compared with controls (2.8±0.61, 2.7±0.55, and 2.7±0.71 vs. 3.6±0.56 L/min/m², respectively; p<0.001 for all comparisons), with no significant difference across MR severity groups (p=0.15).

No LGE was observed in controls with available LGE imaging (15 out of 20 controls) (Table 1). In contrast, LGE prevalence increased stepwise with MR severity, affecting 17% of patients with mild, 46% with moderate, and 51% with severe MR. Compared with controls, LGE prevalence was significantly higher in patients with moderate (p=0.001) and severe MR (p<0.001) (Figure 1). Global native T1 was higher in MVP patients across all severity groups, with significantly higher values in mild, moderate and severe MR compared with controls (1022.0±34.7 ms, 1019.1±28.9 ms and 1026.1±32.5 ms vs. 982.6±34.1 ms, respectively; p<0.001 for all comparisons) (Figure 2). Global ECV was similar between controls and MR groups, whereas inferolateral ECV was elevated in severe MR (28.7±4.8% vs. 25.8±2.9% in controls, p=0.03) and increased with MR severity (p=0.01). Furthermore, iECV and iICV were elevated in patients with severe MR compared with controls and increased with MR severity (Figure 3).

**Figure 1.**
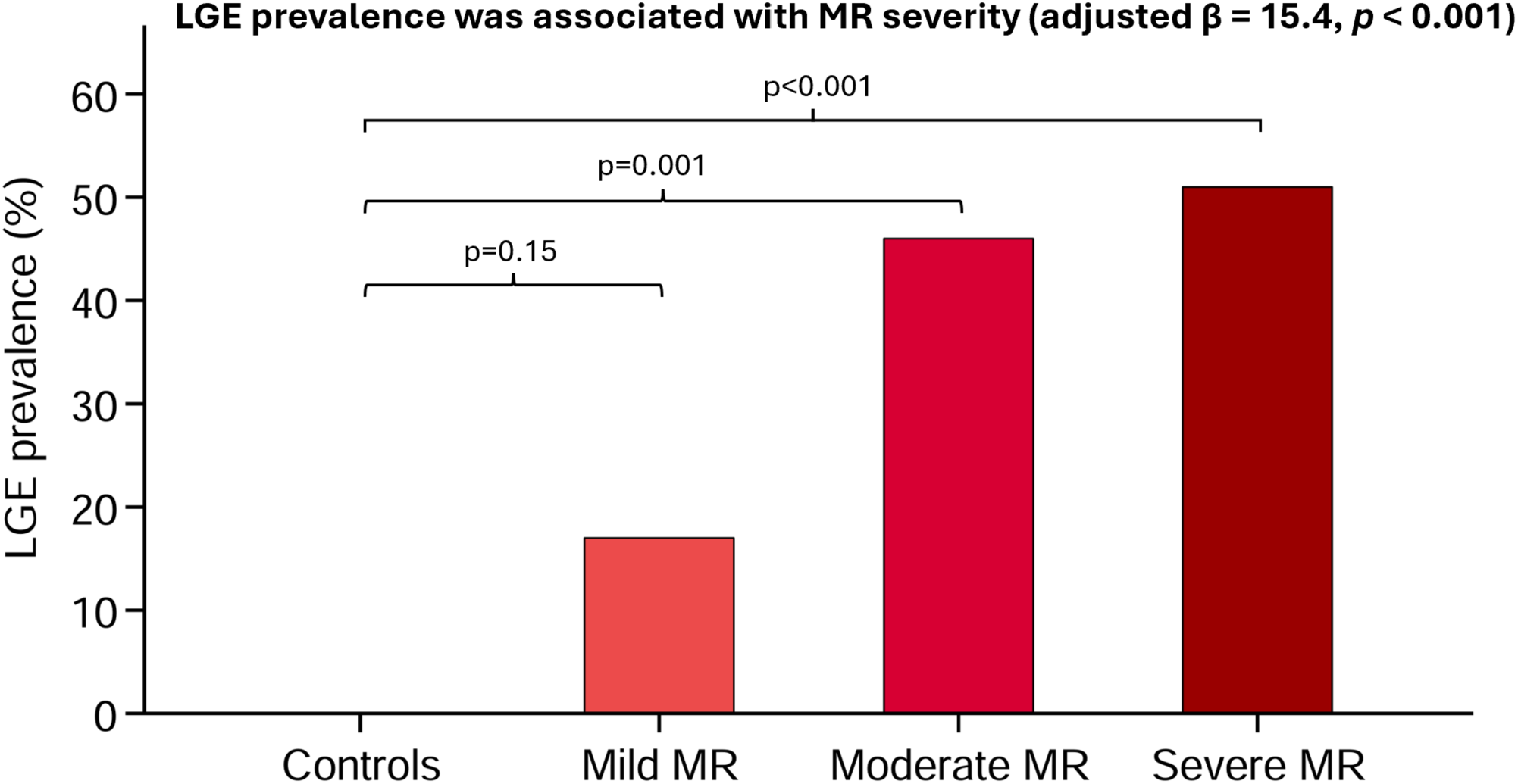
LGE prevalence across categories of MR severity in patients with MVP compared to controls. Bar graph shows percentage of patients with LGE in controls and patients stratified by MR severity (mild <20%, moderate 20-39%, severe ≥40% MR fraction). Unstandardized β-coefficient from adjusted linear regression with MR fraction as continuous variable.

**Figure 2.**
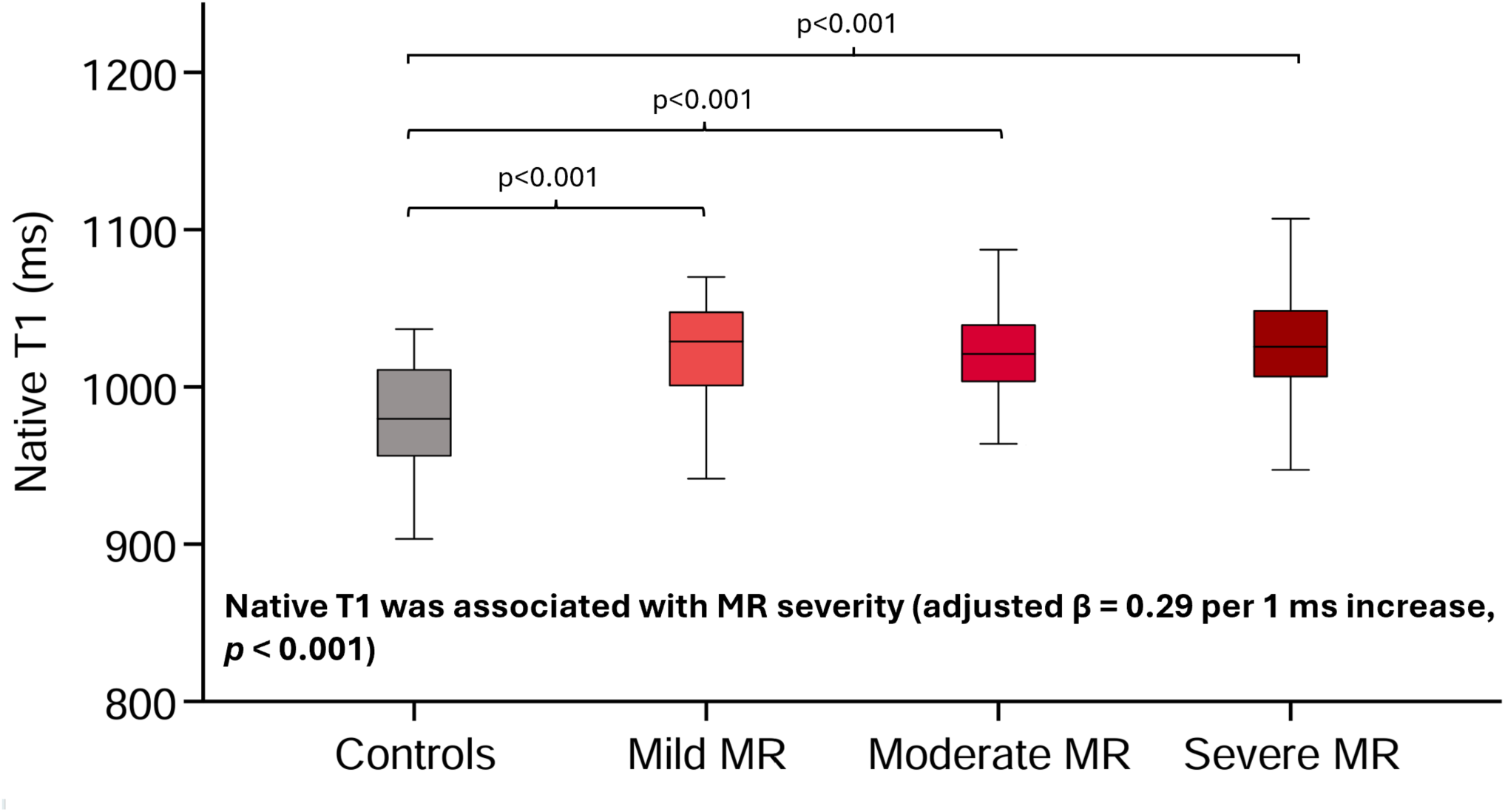
Global native T1 values across categories of MR severity in patients with MVP compared to controls. Box plots show median and interquartile range for global native T1 in controls and patients stratified by MR severity (mild <20%, moderate 20-39%, severe ≥40% MR fraction). Unstandardized β-coefficient from adjusted linear regression with MR fraction as continuous variable.

**Figure 3.**
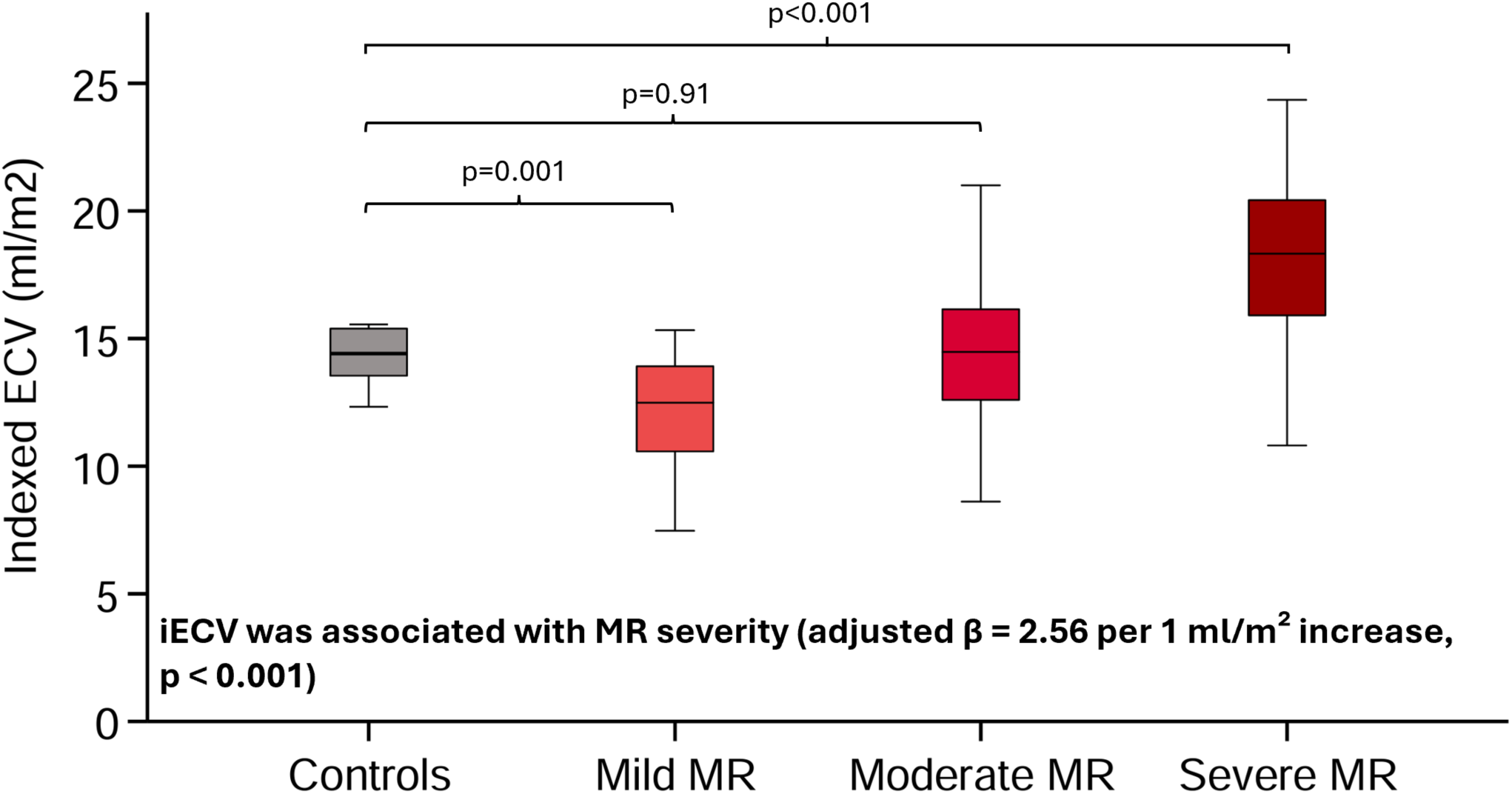
Indexed ECV across categories of MR severity in patients with MVP compared to controls. Box plots show median and interquartile range for indexed ECV in controls and patients stratified by MR severity (mild <20%, moderate 20-39%, severe ≥40% MR fraction). Unstandardized β-coefficient from adjusted linear regression with MR fraction as continuous variable.

### Characteristics Associated with Increasing MR Severity

As shown in Table 2, in crude analyses the characteristics most strongly associated with increasing MR severity were presence of LGE (β 9.87, p=0.002), iECV (β 2.59, p<0.001), iICV (β 0.95, p<0.001), EF (β 0.74, p<0.001), LVMI (β 0.71, p<0.001), ESVI (β 0.36, p<0.001), EDVI (β 0.35, p<0.001) and age (β 0.21, p=0.04). Female sex (β −10.7, p<0.001) and cardiac index (β –7.01, p=0.01) were associated with lower MR severity (Table 2). After adjustment, all these characteristics, except for age and female sex, remained significantly associated with MR severity. EDV/LVM-ratio and global native T1 were not significantly associated with MR severity in crude analyses, but both became significantly associated after adjustment (β 27.1, p<0.001 and β 0.29, p<0.001, respectively).

**Table 2.** Characteristics associated with MR severity* in patients with MVP.

|  | Unadjusted |  | Adjusted** |  |
| --- | --- | --- | --- | --- |
| | $\beta$ (95% CI) | p-value | $\beta$ (95% CI) | p-value |
| **Clinical variables** |  |  |  |  |
| Age, years | 0.21 (0.01, 0.40) | 0.04 |  |  |
| Sex, female | -10.7 (-16.6, -4.70) | <0.001 |  |  |
| BMI, kg/m<sup>2</sup> | 0.27 (-0.62, 1.15) | 0.55 |  |  |
| Hematocrit, % | -0.28 (-0.96, 0.40) | 0.41 |  |  |
| Diabetes | 4.73 (-19.1, 28.5) | 0.70 |  |  |
| Hypertension | 3.03 (-4.75, 10.8) | 0.44 |  |  |
| Atrial fibrillation | 7.32 (-0.37, 15.0) | 0.06 |  |  |
| Hyperlipidemia | -0.10 (-12.3, 12.2) | 0.99 |  |  |
| NTproBNP, pg/ml | 0.004 (0.00, 0.01) | 0.08 | 0.01 (0.00, 0.01) | 0.06 |
| **Mitral valve characteristics** |  |  |  |  |
| Barlow's disease | -4.37 (-10.6, 1.87) | 0.17 | -0.77 (-7.27, 5.74) | 0.82 |
| Other etiology | 4.37 (-1.87, 10.6) | 0.17 | 0.77 (-5.74, 7.27) | 0.82 |
| Posterior leaflet prolapse | 8.28 (2.19, 14.4) | 0.01 | 4.17 (-2.65, 11.0) | 0.23 |
| Anterior leaflet prolapse | -5.56 (-20.8, 9.69) | 0.47 | -2.91 (-17.8, 11.9) | 0.70 |
| Bileaflet prolapse | -7.39 (-13.5, -1.24) | 0.02 | -3.36 (-10.0, 3.29) | 0.32 |
| MAD inferolateral | -6.16 (-13.5, 1.22) | 0.10 | -3.61 (-11.1, 3.91) | 0.34 |
| MAD inferior | -4.17 (-11.5, 3.16) | 0.26 | -2.00 (-9.30, 5.29) | 0.59 |
| **CMR measures** |  |  |  |  |
| EF, % | 0.74 (0.34, 1.13) | <0.001 | 0.89 (0.44, 1.33) | <0.001 |
| EDVI, ml/m<sup>2</sup> | 0.35 (0.26, 0.44) | <0.001 | 0.45 (0.35, 0.56) | <0.001 |
| ESVI, ml/m<sup>2</sup> | 0.36 (0.17, 0.56) | <0.001 | 0.44 (0.21, 0.67) | <0.001 |
| LVMI, g/m<sup>2</sup> | 0.71 (0.52, 0.89) | <0.001 | 0.86 (0.53, 1.19) | <0.001 |
| Cardiac index, l/min/ m<sup>2</sup> | -7.01 (-12.3, -1.76) | 0.01 | -7.91 (-13.8, -1.98) | 0.01 |
| EDV/LVM-ratio | 4.26 (-5.30, 13.8) | 0.38 | 27.1 (18.0, 36.1) | <0.001 |
| LGE prevalence | 9.87 (3.67, 16.1) | 0.002 | 15.4 (7.77, 23.0) | <0.001 |
| LGE, ml | 2.14 (0.79, 3.49) | 0.002 | 2.95 (1.27, 4.64) | <0.001 |
| Global native T1, ms | 0.07 (-0.03, 0.18) | 0.15 | 0.29 (0.18, 0.39) | <0.001 |
| Global native T2, ms | -0.56 (-2.13, 1.01) | 0.48 | 0.13 (-1.65, 1.90) | 0.89 |
| Global ECV, % | 0.85 (-0.39, 2.08) | 0.18 | 1.19 (-0.30, 2.67) | 0.12 |
| ECV inferolateral, % | 1.06 (0.30, 1.83) | 0.01 | 1.30 (0.37, 2.24) | 0.01 |
| iECV, ml/m<sup>2</sup> | 2.59 (1.90, 3.28) | <0.001 | 2.56 (1.47, 3.64) | <0.001 |
| iICV, ml/m<sup>2</sup> | 0.95 (0.67, 1.22) | <0.001 | 1.19 (0.69, 1.68) | <0.001 |

\*Reflected by MR fraction as a continuous variable in the model
\*\*Adjusted for age, sex, body mass index and hypertension.
Abbreviations: BMI – body mass index, ECV – extracellular volume, EDVI – end diastolic volume index, EF – ejection fraction, ESVI – end systolic volume index, LGE – late gadolinium enhancement, LV – left ventricular, LVMI – left ventricular mass index, iECV – indexed extracellular volume, iICV – indexed intracellular volume, MAD – mitral annular disjunction, MR – mitral regurgitation, MVP – mitral valve prolapse, NYHA – New York Heart Association.

### Characteristics Associated with LV Dilatation

As presented in Table 3, univariable analyses showed that the characteristics most strongly associated with LV dilatation were iECV (OR 1.42, p<0.001), ESVI (OR 1.15, p<0.001), iICV (OR 1.15, p<0.001), LVMI (OR 1.11, p<0.001), MR fraction (OR 1.05, p<0.001) and MR volume (OR 1.05, p<0.001). In adjusted analyses, the strongest independent predictors of LV dilatation were iECV (OR 1.59, p<0.001), iICV (OR 1.43, p<0.001), LVMI (OR 1.23, p<0.001), ESVI (OR 1.18, p<0.001), MR fraction (OR 1.06, p<0.001), MR volume (OR 1.06, p<0.001) and global native T1 (OR 1.02, p=0.04). No significant associations were observed for age, comorbidities, or specific mitral valve characteristic subtypes.

**Table 3.** Clinical and CMR characteristics associated with LV dilatation in patients with MVP.

|  | Unadjusted |  | Adjusted* |  |
| --- | --- | --- | --- | --- |
|  | Odds ratio (95% CI) | p-value | Odds ratio (95% CI) | p-value |
| **Clinical variables** |  |  |  |  |
| Age, years | 1.00 (0.97-1.03) | 0.94 |  |  |
| Sex, female | 0.74 (0.33-1.67) | 0.47 |  |  |
| BMI, kg/m<sup>2</sup> | 1.00 (0.89-1.12) | 1.00 |  |  |
| Hematocrit, % | 0.88 (0.79-0.97) | 0.02 |  |  |
| Hypertension | 1.71 (0.58-5.07) | 0.33 |  |  |
| Atrial fibrillation | 0.98 (0.36-2.65) | 0.97 |  |  |
| Hyperlipidemia | 1.32 (0.25-6.87) | 0.75 |  |  |
| NTproBNP, pg/ml | 1.000 (0.999-1.000) | 0.26 | 0.999 (0.999-1.000) | 0.17 |
| **Mitral valve characteristics** |  |  |  |  |
| Barlow's disease | 1.04 (0.47-2.33) | 0.92 | 1.18 (0.49-2.84) | 0.71 |
| Other etiology | 0.96 (0.43-2.15) | 0.92 | 0.85 (0.35-2.03) | 0.71 |
| Posterior leaflet prolapse | 1.37 (0.61-3.07) | 0.45 | 1.36 (0.54-3.43) | 0.51 |
| Anterior leaflet prolapse | 0.27 (0.04-1.69) | 0.27 | 0.22 (0.03-1.48) | 0.12 |
| Bileaflet prolapse | 0.94 (0.42-2.11) | 0.88 | 1.04 (0.43-2.55) | 0.93 |
| MAD inferolateral | 1.58 (0.57-4.38) | 0.38 | 1.75 (0.60-5.13) | 0.31 |
| MAD inferior | 1.05 (0.41-2.70) | 0.93 | 1.12 (0.42-3.00) | 0.83 |
| **CMR measures** |  |  |  |  |
| MR fraction, % | 1.05 (1.02-1.09) | <0.001 | 1.06 (1.03-1.09) | <0.001 |
| MR volume, ml | 1.05 (1.02-1.07) | <0.001 | 1.06 (1.03-1.08) | <0.001 |
| EF, % | 1.01 (0.96-1.07) | 0.66 | 1.02 (0.96-1.07) | 0.60 |
| ESVI, ml/m<sup>2</sup> | 1.15 (1.08-1.21) | <0.001 | 1.18 (1.09-1.26) | <0.001 |
| LVMI, g/m<sup>2</sup> | 1.11 (1.05-1.16) | <0.001 | 1.23 (1.13-1.34) | <0.001 |
| Cardiac index, l/min/ m<sup>2</sup> | 1.60 (0.80-3.21) | 0.18 | 1.83 (0.80-4.18) | 0.15 |
| LGE prevalence | 2.11 (0.89-4.99) | 0.09 | 2.38 (0.90-6.33) | 0.08 |
| LGE, ml | 1.15 (0.93-1.41) | 0.20 | 1.17 (0.93-1.48) | 0.19 |
| Global native T1, ms | 1.01 (1.00-1.03) | 0.09 | 1.02 (1.00-1.03) | 0.04 |
| Global native T2, ms | 1.01 (0.82-1.25) | 0.93 | 0.98 (0.79-1.23) | 0.87 |
| Global ECV, % | 1.08 (0.92-1.28) | 0.34 | 1.11 (0.93-1.33) | 0.25 |
| ECV inferolateral, % | 1.09 (0.96-1.23) | 0.18 | 1.07 (0.94-1.21) | 0.31 |
| iECV, ml/m<sup>2</sup> | 1.42 (1.20-1.69) | <0.001 | 1.59 (1.29-1.97) | <0.001 |
| iICV, ml/m<sup>2</sup> | 1.15 (1.08-1.23) | <0.001 | 1.43 (1.23-1.66) | <0.001 |

\*Adjusted for age, sex, body mass index and hypertension.
Abbreviations: BMI – body mass index, ECV – extracellular volume, EDVI – end diastolic volume index, EF – ejection fraction, ESVI – end systolic volume index, LGE – late gadolinium enhancement, LV – left ventricular, LVMI – left ventricular mass index, iECV – indexed extracellular volume, iICV – indexed intracellular volume, MAD – mitral annular disjunction, MVP – mitral valve prolapse, NYHA – New York Heart Association.

### Characteristics Associated with Functional Status in Patients with MR

As shown in Table 4, in crude analyses the strongest associations with NYHA class ≥ II were observed for iECV (OR 1.15, p=0.01), followed by age (OR 1.04, p=0.01), iICV (OR 1.04, p=0.04), MR fraction (OR 1.03, p=0.01), LVMI (OR 1.04, p=0.02), and EDV/LVM-ratio (OR 0.15, p=0.004). In adjusted analyses, higher global native T1 (OR 1.02, p=0.004), iECV (OR 1.15, p=0.04) and lower EDV/LVM-ratio (OR 0.16, p=0.02) remained associated with higher NYHA class. In a subgroup analysis restricted to patients with MVP referred for mitral valve surgery, global native T1 was the only CMR parameter that differed significantly between asymptomatic patients (NYHA class I, n=24) and those with symptoms (NYHA class ≥ II, n=40) (1007.2±25.9 ms vs. 1028.0±26.6 ms, p=0.003), despite comparable MR fraction, LV volumes, LGE prevalence and iECV (Table 5).

**Table 4.** Clinical and CMR characteristics associated with symptoms (NYHA class ≥ II) in patients with MVP.

|  | Unadjusted |  | Adjusted* |  |
| --- | --- | --- | --- | --- |
|  | Odds ratio (95% CI) | p-value | Odds ratio (95% CI) | p-value |
| **Clinical variables** |  |  |  |  |
| Age, years | 1.04 (1.01-1.06) | 0.01 |  |  |
| Sex, female | 0.55 (0.26-1.17) | 0.12 |  |  |
| BMI, kg/m<sup>2</sup> | 1.05 (0.95-1.17) | 0.36 |  |  |
| Hematocrit, % | 1.02 (0.94, 1.10) | 0.69 |  |  |
| Diabetes | 1.12 (0.07-18.3) | 0.94 |  |  |
| Hypertension | 1.99 (0.78-5.09) | 0.15 |  |  |
| Atrial fibrillation | 1.99 (0.78-5.09) | 0.15 |  |  |
| Hyperlipidemia | 0.65 (0.15-2.85) | 0.57 |  |  |
| NTproBNP, pg/ml | 1.001 (1.000-1.002) | 0.07 | 1.001 (1.000-1.002) | 0.17 |
| **Mitral valve characteristics** |  |  |  |  |
| Barlow's disease | 0.99 (0.47-2.08) | 0.98 | 1.62 (0.69-3.82) | 0.27 |
| Other etiology | 1.01 (0.48-2.13) | 0.98 | 0.62 (0.26-1.45) | 0.27 |
| Posterior leaflet prolapse | 1.24 (0.59-2.61) | 0.57 | 0.67 (0.27-1.62) | 0.37 |
| Anterior leaflet prolapse | 1.71 (0.28-10.7) | 0.57 | 3.15 (0.44-22.7) | 0.25 |
| Bileaflet prolapse | 0.73 (0.35-1.55) | 0.42 | 1.18 (0.50-2.77) | 0.71 |
| MAD inferolateral | 0.51 (0.20-1.26) | 0.14 | 0.77 (0.29-2.07) | 0.60 |
| MAD inferior | 0.58 (0.24-1.40) | 0.22 | 0.79 (0.31-2.03) | 0.62 |
| **CMR measures** |  |  |  |  |
| MR fraction, % | 1.03 (1.01-1.06) | 0.01 | 1.03 (1.00-1.05) | 0.06 |
| MR volume, ml | 1.01 (1.00-1.02) | 0.12 | 1.01 (0.99-1.02) | 0.33 |
| EF, % | 1.02 (0.97-1.07) | 0.47 | 1.02 (0.97-1.07) | 0.52 |
| EDVI, ml/m<sup>2</sup> | 1.00 (0.99-1.01) | 0.97 | 0.997 (0.98-1.01) | 0.71 |
| ESVI, ml/m<sup>2</sup> | 1.00 (0.98-1.02) | 0.94 | 0.99 (0.97-1.02) | 0.68 |
| LVMI, g/m<sup>2</sup> | 1.04 (1.01-1.07) | 0.02 | 1.04 (1.00-1.09) | 0.05 |
| EDV/LVM-ratio | 0.15 (0.04-0.54) | 0.004 | 0.16 (0.04-0.71) | 0.02 |
| Cardiac index, l/min/ m<sup>2</sup> | 0.57 (0.30-1.09) | 0.09 | 0.74 (0.35-1.58) | 0.44 |
| LGE prevalence | 1.63 (0.76-3.50) | 0.21 | 1.12 (0.47-2.70) | 0.80 |
| LGE, ml | 1.17 (0.98-1.40) | 0.08 | 1.09 (0.90-1.33) | 0.37 |
| Global native T1, ms | 1.01 (1.00-1.02) | 0.10 | 1.02 (1.01-1.04) | 0.004 |
| Global native T2, ms | 0.90 (0.74-1.08) | 0.24 | 0.81 (0.64-1.02) | 0.07 |
| Global ECV, % | 1.08 (0.93-1.25) | 0.31 | 1.10 (0.94-1.30) | 0.25 |
| ECV inferolateral, % | 1.04 (0.95-1.15) | 0.38 | 1.01 (0.92-1.12) | 0.82 |
| iECV, ml/m<sup>2</sup> | 1.15 (1.03-1.28) | 0.01 | 1.15 (1.01-1.32) | 0.04 |
| iICV, ml/m<sup>2</sup> | 1.04 (1.00-1.09) | 0.04 | 1.06 (0.995-1.12) | 0.07 |

\*Adjusted for age, sex, body mass index and hypertension.
Abbreviations: BMI – body mass index, ECV – extracellular volume, EDVI – end diastolic volume index, EF – ejection fraction, ESVI – end systolic volume index, LGE – late gadolinium enhancement, LV – left ventricular, LVMI – left ventricular mass index, iECV – indexed extracellular volume, iICV – indexed intracellular volume, MAD – mitral annular disjunction, MVP – mitral valve prolapse, NYHA – New York Heart Association.

**Table 5.** Clinical and CMR characteristics of MVP patients referred for mitral valve surgery stratified by NYHA class.

|  | **NYHA-class I**<br>**n=24** | **NYHA-class ≥ II**<br>**n=40** | **p-value** |
| --- | --- | --- | --- |
| **Clinical variables** |  |  |  |
| Age, years | 59.0±14.1 | 58.3±13.9 | 0.81 |
| Sex, female | 25% (6) | 30% (12) | 0.67 |
| BMI, kg/m<sup>2</sup> | 24.9±3.1 | 24.1±4.1 | 0.38 |
| Hematocrit, % | 42.0±3.6 | 42.6±3.1 | 0.49 |
| Diabetes | 8% (2) | 3% (1) | 0.55 |
| Hypertension | 29% (7) | 30% (12) | 0.94 |
| Atrial fibrillation | 21% (5) | 28% (11) | 0.55 |
| Hyperlipidemia | 21% (5) | 8% (3) | 0.14 |
| NTproBNP, pg/ml | 314.6±374.5 | 540.4±782.2 | 0.50 |
| **Mitral valve characteristics** |  |  |  |
| Barlow's disease | 42% (10) | 43% (17) | 0.95 |
| Other etiology | 58% (14) | 58% (23) | 0.95 |
| Posterior leaflet prolapse | 71% (17) | 65% (26) | 0.63 |
| Anterior leaflet prolapse | 8% (2) | 3% (1) | 0.55 |
| Bileaflet prolapse | 21% (5) | 33% (13) | 0.32 |
| MAD inferolateral | 4% (1) | 13% (5) | 0.40 |
| MAD inferior | 13% (3) | 13% (5) | 1.00 |
| **CMR measures** |  |  |  |
| MR fraction, % | 43.7±13.6 | 41.6±12.9 | 0.53 |
| MR volume, ml | 72.2±37.8 | 61.1±28.4 | 0.23 |
| EF, % | 60.6±6.9 | 59.5±6.8 | 0.52 |
| LV dilatation | 92% (22) | 78% (31) | 0.19 |
| EDVI, ml/m<sup>2</sup> | 135.1±32.1 | 121.5±27.9 | 0.11 |
| ESVI, ml/m<sup>2</sup> | 54.3±20.5 | 49.5±15.3 | 0.56 |
| LVMI, g/m<sup>2</sup> | 68.7±12.6 | 66.9±13.6 | 0.62 |
| EDV/LVM-ratio | 2.08±0.26 | 1.93±0.36 | 0.10 |
| Cardiac index, l/min/ m<sup>2</sup> | 2.67±0.47 | 2.66±0.58 | 0.95 |
| LGE prevalence | 54% (13) | 55% (22) | 0.95 |
| LGE, ml | 2.1±2.4 | 2.3±2.8 | 1.00 |
| Global native T1, ms | 1007.2±25.9 | 1028.0±26.6 | 0.003 |
| Global native T2, ms | 49.4±2.3 | 48.8±2.2 | 0.29 |
| Global ECV, % | 26.9±1.6 | 27.4±3.1 | 0.45 |
| ECV inferolateral, % | 28.1±4.7 | 28.4±4.5 | 0.83 |
| iECV, ml/m<sup>2</sup> | 17.6±3.0 | 17.4±3.8 | 0.92 |
| iICV, ml/m<sup>2</sup> | 47.8±9.2 | 49.3±10.0 | 0.57 |

Abbreviations: BMI – body mass index, ECV – extracellular volume, EDVI – end diastolic volume index, EF – ejection fraction, ESVI – end systolic volume index, LGE – late gadolinium enhancement, LV – left ventricular, LVMI – left ventricular mass index, iECV – indexed extracellular volume, iICV – indexed intracellular volume, MAD – mitral annular disjunction, MVP – mitral valve prolapse, MR – mitral regurgitation, NYHA – New York Heart Association.

## Discussion

There were several key findings from this comprehensive analysis of the components and extent of LV remodelling across the spectrum of MR severity in MVP, and their associations with functional status. First, iECV increased stepwise with MR severity, paralleled by a marked increase in LGE prevalence (0% in controls, 17% in mild, 46% in moderate, and 51% in severe MR) and the prevalence of LV dilatation (15%, 46%, 67%, and 88%, respectively). Second, in adjusted analyses, presence of LGE, higher EDV/LVM-ratio, higher EDVI, elevated global native T1 and iECV were the variables most strongly associated with greater MR severity. Third, in crude analyses, higher MR fraction, LVMI, iECV, and a lower EDV/LVM-ratio were associated with higher NYHA class, whereas in adjusted models only the EDV/LVM-ratio, iECV and global native T1 remained significant.

Despite increasing recognition of CMR-derived markers of LV remodelling in MVP, data describing the presence, components and extent of myocardial remodelling across different stages of MR in MVP, and how these changes relate to functional status, remain limited. Prior studies linking MR severity with CMR tissue characteristics have mainly focused on focal fibrosis detected by LGE (19, 20), diffuse myocardial fibrosis derived from regional rather than global measurements (21–23) or reporting only post-contrast T1 values (22). Many of these studies are further limited by modest sample sizes (n=34, 41, 73) (21, 22, 24), restriction to moderate-to-severe MR (24), heterogeneous cohorts that mix patients with and without prior myocardial infarction (21), exclusion of patients with reduced EF or significant concentric LV hypertrophy (23), or the absence of healthy controls (19, 20, 23). Moreover, symptom burden has often been inferred indirectly from hard clinical endpoints such as mitral valve surgery (23), ventricular arrhythmias (19–22), new-onset atrial fibrillation (19), arterial embolism (19), heart failure (19), or cardiovascular death (19, 23), thereby capturing functional status mainly in advanced disease. To date, the present study is the first to simultaneously assess absolute myocardial compartment volumes (iECV and iICV), global diffuse myocardial tissue characteristics (ECV and native T1/T2), focal fibrosis (LGE) and patient-reported functional status by NYHA class across the full spectrum of MR severity.

### Myocardial Tissue Characteristics Across MR Severity

Previous CMR studies have demonstrated a high prevalence of myocardial involvement in patients with MVP and MR using preliminary LGE to identify focal fibrosis (19, 20). An association between MR severity and ECV has also been reported, but ECV was measured only from regional mid-ventricular septal segments that excluded LGE (23). While midventricular septal ECV is often used as a surrogate of diffuse myocardial fibrosis (2), this approach may be limited in conditions such as MVP, where regional heterogeneity in myocardial remodelling is common (23). Global ECV may therefore differ when all LV segments, including those containing non-ischaemic LGE, are incorporated. In contrast, a study restricted to only severe MR used global LV assessment based on basal and midventricular short-axis segmentations, including non-infarction LGE, and showed that MR severity was related to higher iECV and iICV but not ECV (16), findings that support the observations in the present study. By additionally incorporating apical short-axis segmentations, our methodology provides a more comprehensive assessment of global myocardial involvement than approaches limited to mid-septal regions or basal and midventricular levels alone.

With respect to T1 mapping, one earlier study reported a borderline association between MR severity, quantified by MR fraction and post-contrast T1 (22), whereas two others found no such associations (21, 24). These discrepancies likely reflect limited sample sizes (21, 22, 24), a focus on only moderate-to-severe MR (24), inclusion of patients with prior myocardial infarction (21), use of regional mapping approaches (21, 22), and reporting post-contrast but not native T1 values (22). In another previous study (n=34), patients with MVP and MAD had higher native T2, indicative of subclinical myocardial edema (25), patients with hemodynamically significant MR were not included, why the distribution of native T2 across the spectrum of MR severity could not be assessed (25).

In the present study, the prevalence and extent of LGE increased progressively with MR severity, likely reflecting the systolic stretch of the myocardium from the prolapsing leaflets and elongated chordae (19, 21). In parallel, iECV and global native T1 rose with MR severity, indicating diffuse interstitial expansion in the setting of worsened hemodynamic burden. iICV, which isolates the myocyte compartment and reflects cardiomyocyte hypertrophy, also increased with increasing MR severity. The concordant rise in iICV, iECV, and native T1 therefore points to combined cardiomyocyte hypertrophy and interstitial expansion as key components of LV remodelling in MVP-related MR. Importantly, these CMR markers of maladaptive remodelling were already abnormal in mild and moderate MR, indicating that myocardial injury or structural alterations may develop in parallel with, or even precede, the most advanced stages of MR.

### LV Remodelling and Functional Status

Symptom burden in MVP appears to reflect underlying myocardial remodelling more closely than MR severity alone. In the present cohort, patients with NYHA class ≥ II had a greater LGE burden, higher iECV, iICV, global native T1, MR fraction and LVMI, and a lower EDV/LVM-ratio compared with asymptomatic patients. In adjusted models, only global native T1, iECV and EDV/LVM-ratio remained independently associated with NYHA class. These findings suggest that the transition from compensated to symptomatic MR may be driven by progressive interstitial remodelling, even before overt systolic dysfunction or severe regurgitation develops.

The inverse relationship between EDV/LVM-ratio and symptom burden further supports the concept that disproportionate hypertrophic remodelling, rather than chamber dilation per se, contributes to more severe symptoms, implying that CMR-derived markers of hypertrophy and fibrosis may help identify patients at risk of clinical deterioration before conventional MR thresholds are met. We found one previous study that has examined the relationship between myocardial tissue characteristics and NYHA class, reporting borderline higher ECV in symptomatic (NYHA class ≥ II) patients with primary MR, while native T1 did not reach statistical significance (26). In that cohort, only surgical patients with severe MR were included (26), whereas our study spans the whole spectrum of MR severity, potentially allowing native T1 to capture cellular and interstitial changes before extracellular volume expansion becomes apparent. These observations suggest that native T1 may be a sensitive marker of early interstitial remodelling linked to symptom development.

In a subgroup analysis restricted to patients with severe MR, native T1 was the only CMR parameter that differed significantly between asymptomatic patients and those with symptoms, despite comparable MR fraction, LV volumes and LGE prevalence. This finding suggests that native T1 is a versatile biomarker of diffuse myocardial remodelling, which may reflect a transition toward symptomatic disease independently of the degree of volume overload, and raises the hypothesis that elevated native T1 could serve as an imaging threshold to guide earlier surgical intervention in patients with severe MR before overt LV dysfunction develops. In contrast, native T2 was not associated with symptoms, consistent with its specificity for acute inflammation and edema (7).

## Limitations

Patients with mild or moderate MR underwent CMR in a clinical context, either as part of the assessment of mitral valve disease (MVP, MR, and MAD), which may limit generalizability to an unselected MVP population. Moreover, focal fibrosis was assessed only in the LV myocardium, and not in the papillary muscles, because dark-blood LGE acquisitions, required for optimal papillary muscle imaging, were unavailable in the majority of the examinations. This may have resulted in an underestimation of the overall focal fibrosis burden in patients with MVP. Finally, the cross-sectional design precludes conclusions about causal relationships between MR severity, CMR markers of LV remodelling, and symptom burden.

## Conclusion

In patients with MVP, CMR markers of maladaptive LV remodelling were abnormal already in the early stages of MR and increased with MR severity. EDV/LVM-ratio, iECV and global native T1 showed the strongest associations with symptom burden, rather than MR severity alone. These findings suggest that CMR-derived markers of LV hypertrophy and fibrosis may help identify patients at risk of clinical deterioration before conventional MR thresholds for surgery are reached.

## Data Availability

The data underlying this article will be shared on reasonable request to the corresponding author.

## Clinical Perspectives

### COMPETENCY IN MEDICAL KNOWLEDGE

In patients with MVP, CMR-derived markers of LV remodelling are already abnormal in early MR and progress with increasing MR severity, with native T1, iECV, iICV, and EDV/LVM-ratio capturing diffuse interstitial expansion and hypertrophic response beyond conventional volumetric measures. EDV/LVM-ratio, iECV and global native T1 showed the closest relationship with symptom burden.

### TRANSLATIONAL OUTLOOK

Integrating quantitative CMR markers of LV remodelling into routine assessment of MVP could help identify patients with early myocardial injury who might benefit from closer follow-up or earlier valve intervention, despite only mild-to-moderate MR. Future longitudinal studies are needed to evaluate management strategies that incorporate thresholds for native T1, iECV and EDV/LVM-ratio, to determine whether CMR-guided timing of surgery improves symptoms, promotes reverse remodelling, and reduces adverse outcomes.

## Acknowledgement

The authors wish to acknowledge Kelvin Chow at Siemens Healthcare Ltd, Calgary, AB, Canada, for providing the research sequence for native T1 and ECV mapping.

## Abbreviations

CMR: Cardiovascular magnetic resonance
ECV: Extracellular volume fraction
EDVI: End-diastolic volume index
EF: Ejection fraction
ESVI: End-systolic volume index
FED: Fibroelastic deficiency
iECV: Indexed extracellular volume
iICV: Indexed intracellular volume
LA: Left atrium
LGE: Late gadolinium enhancement
LV: Left ventricle
LVMI: Left ventricular mass index
MAD: Mitral annular disjunction
MR: Mitral regurgitation
MVP: Mitral valve prolapse
NYHA: New York Heart Association functional class

## Notes

**Conflicts of Interest:** Nothing to Disclose.

### Competing Interest Statement

The authors have declared no competing interest.

### Clinical Trial

Clinical Trial Registration: NCT06255457

### Author Declarations

The study was approved by the Swedish Ethical Review Authority (Etikprövningsmyndigheten), Sweden (Dnr 2023-01904-01).

